# Health facility support and its influence on weight gain for neonates under Kangaroo Mother Care in Western Kenya

**DOI:** 10.1101/2025.02.05.25321723

**Authors:** Seline Mukabi, Everlyne Nyanchera Morema, Mary Kipmerewo, Morris Senghor Shisanya

**Affiliations:** Department of Reproductive Health, Midwifery and Child Health, School of Nursing, Midwifery and Paramedical Sciences (SONMAPS), Masinde Muliro University of Science and Technology (MMUST), Kakamega, Kenya; Department of Community Health Nursing, School of Nursing, Kibabii University, Bungoma, Kenya

**Keywords:** Kangaroo Mother Care, low Birthweight, facility related factors

## Abstract

**Introduction:** Kangaroo Mother Care (KMC) is a recommended cost-effective intervention for low birthweight neonates that promotes neonatal growth, and survival. Its effectiveness is dependent on several factors. This study examines facility factors that influence outcome of KMC in Western Kenya.

**Methods:** This was a mixed methods study that examined facility support factors influencing KMC outcomes in Western Kenya. A multi-stage sampling approach was used attain the sample size (275). Data collection involved structured questionnaires for caregivers and key informant interviews (KIIs) for healthcare providers. Quantitative data was analyzed using SPSS v26. Binary logistic regression used to determine influence of facility support on KMC neonates weight gain with α = 0.05. Qualitative data from 12 KIIs was thematically analyzed to provide deeper insights into the factors affecting KMC implementation.

**Results:** The average facility KMC implementation score was 10.78±2.59 out of 19 (56.71%±13.65%). Only 38.18% of neonates achieved the recommended daily weight gain (≥15g/kg). Average daily weight gain was 12.96g±4.85g. The average length of stay on KMC was 36.25±10.09 days, less than the average recommended 41.21±16.02 days for optimal weight gain. Facility-related predictors of achieving ≥15g/kg daily weight gain included the availability of breast pumps (AOR:2.81, P=0.009), milk banks (AOR:3.22, P=0.004), adequate food for mothers (AOR:5.32, P<0.001), seating for mothers (AOR:3.72, P=0.001), dedicated rooms for breast milk expression (AOR:4.77, P<0.001), presence of easily understandable KMC information (AOR:2.86, P=0.007), family-centered KMC support (AOR:2.41, P=0.032), and adequate staffing for KMC (AOR:3.44, P=0.002). The qualitative data showed that nurses played key roles in clinical care, education, research, advocacy, and community engagement for KMC. While formal training improved competency, gaps remained, emphasizing the need for continuous learning..

**Conclusions:** Facility level support is crucial for maximizing benefits of KMC especially through provision of maternal nutrition, infrastructure support, guaranteeing maternal comfort, family-centered care and adequate staffing.

## Introduction

Kangaroo Mother Care is a vital intervention for low birth weight (LBW) infants, involving prolonged skin-to-skin contact between the infant and mother or caregiver. This low-cost alternative to traditional neonatal care, which typically relies on incubators and specialized staff, has been shown to reduce neonatal mortality and morbidity. KMC promotes growth, enhances weight gain, lowers infection risks, and strengthens the emotional bond between mother and infant [1, 2]. The World Health Organization (WHO) endorses KMC, especially in low-resource settings where incubators are scarce [3]. When practiced for over 8 hours daily, KMC reduces preterm mortality by 40%, improving survival rates for preterm infants [1]. Globally, 15.5% of infants are born with LBW, with the highest rates in low-income countries, where LBW is a major contributor to neonatal deaths [4].

Health facility support factors significantly impact the implementation of KMC for low birth weight infants. These factors include the availability of essential drugs, equipment, staff, health facilities, and financial resources. The availability of drugs is critical, as studies show that hospitals with consistent supplies of antibiotics, vaccines, and nutritional supplements achieve better outcomes in managing LBW infants [5, 6]. Similarly, appropriate equipment like thermometers and weighing scales is necessary for monitoring and managing LBW infants, with hospitals lacking such equipment facing challenges [7]. Adequate staffing is also essential, as trained healthcare providers ensure continuous support for mothers practicing KMC, leading to better neonatal outcomes [8]. Inadequate staffing, as seen in various African studies, can undermine the effectiveness of KMC, leading to higher neonatal morbidity and mortality [9]. Health facility infrastructure also plays a key role; urban hospitals with better infrastructure and dedicated KMC wards report improved neonatal outcomes [10]. Financial resources are another critical factor, enabling the procurement of necessary supplies and ensuring program sustainability. Hospitals with adequate funding tend to have better neonatal outcomes, while those in resource-limited settings struggle with KMC implementation [11]. The prevalence of LBW has been rising, particularly in Busia, Kisii, and Migori counties, where rates range from 18% to 30% [12]. Despite the benefits of KMC, there is limited data on its implementation and outcomes in these regions. This study aims to evaluate the level of health facility KMC support in these counties and its influence on weight gain among neonates as the key indicator of KMC outcome.

## Methods

### Study Design

This study adopted a descriptive and analytical cross-sectional design, incorporating both quantitative (analytical) and qualitative (descriptive) methods to explore factors influencing KMC outcomes. The cross-sectional approach allowed for the assessment of existing conditions and associations at a specific point in time, capturing data on maternal and neonatal care practices in selected health facilities.

### Study area

This study was conducted in Migori, Kisii, and Busia Counties in Western Kenya. These facilities were selected due to their high burden of neonatal mortality associated with low-birth-weight (LBW) babies. The counties have a mix of urban and rural populations and varying levels of healthcare infrastructure, making them ideal for examining facility-related factors influencing KMC outcomes.

The study focused on county referral hospitals and level 4 public health facilities to ensure representation across different healthcare service levels. The study was conducted in Migori, Kisii, and Busia Counties, focusing on key health facilities that provide maternal and neonatal care. In Migori County, the selected study sites included Migori County Referral Hospital, Rongo Subcounty Hospital (Level 4), and Awendo Subcounty Hospital (Level 4). In Kisii County, data was collected from Kisii County Referral Hospital, Gucha District Hospital, and Nyamache Subcounty Hospital. Similarly, in Busia County, the study was conducted at Busia County Referral Hospital, Nambale Subcounty Hospital (Level 4), and Khunyangu Subcounty Hospital (Level 4). These facilities were selected based on availability of KMC services, referral capacity, and their role in maternal and neonatal care. The diverse mix of county referral and sub-county hospitals provided a comprehensive overview of facility-based challenges and enablers of KMC implementation.

### Study Population

The study population comprised mothers and caregivers with low-birth-weight babies at the selected health facilities in Busia, Kisii, and Migori Counties. Additionally, the study included healthcare providers involved in maternity care services at these facilities.

Participants in the study included mothers and caregivers of low-birth-weight infants receiving care at the selected public health facilities. Only those who voluntarily participated and provided written informed consent were included. Additionally, healthcare providers, including doctors, nurses, and registered clinical officers, who were directly involved in maternity service delivery at the study sites, were also part of the study.

The study excluded respondents who were critically ill or suffering from severe medical or surgical conditions at the time of data collection. Individuals below the age of 18 years and those considered mentally unstable or of unsound mind were also not included in the study.

### Sample size determination

The study sample size was determined using the modification of Cochran’s formula, adjusted for finite population correction formula n= (NZ^2^pq)/(e^2^ (N-1)+Z^2^pq), considering a 95% confidence interval (Z = 1.96), a study population (N) of 707, a margin of error (e) of 0.05, and an assumed population proportion (p) of 50% [13]. The calculated sample size was 250, with an additional 10% (25 participants) added to account for non-responses, bringing the final sample to 275 caregivers of low-birth-weight neonates.

Additionally, 12 key informant interviews were conducted with medical officers and nurse-in-charges from the selected health facilities to provide qualitative insights into facility-based factors influencing KMC implementation.

### Sampling Method

The study employed a combination of purposive sampling and random sampling techniques to ensure a representative selection of participants. Purposive sampling was used to select Busia, Kisii, and Migori Counties, based on their high burden of neonatal deaths associated with LBW babies. Within these counties, stratified random sampling was applied to select level 4 public health facilities per county, in addition to purposively including the three county referral hospitals. Proportionate stratification technique was then used to allocate sample sizes per health facility based on their projected workload. This approach ensured that each facility contributed to the sample in proportion to its patient volume, thereby enhancing the generalizability of the findings. Simple random sampling was then employed to select respondent, ensure that every eligible participant had an equal chance of inclusion. This minimized selection bias and increased the validity of the study results.

The final quantitative sample size was 275 caregivers of LBW neonates, distributed across the selected health facilities as follows:

Key informants were purposively selected for participation in KIIs, including medical officers, clinical officers, and nurses delivering maternal and neonatal care services at the selected facilities.

### Data Collection

This study utilized a structured questionnaire to collect quantitative data from mothers and caregivers of LBW babies on KMC. The questionnaire covered demographics, socio- economic factors, institutional influences, and healthcare provider-related factors affecting KMC. It consisted mainly of closed-ended questions for statistical analysis, with optional spaces for qualitative inputs.

For the qualitative component, Key Informant Interviews were conducted with healthcare providers using a structured interview guide. These interviews were audio-recorded (with consent) and supplemented with note-taking. No respondent photos were taken, though facility and equipment images were captured where relevant.

Three trained research assistants, selected from midwifery students, conducted face-to-face interviews using the questionnaire after obtaining written informed consent. Clinical records were reviewed to verify birth weight, gestational age, maternal health history, and neonatal conditions.

The principal investigator conducted KIIs with medical officers, clinical officers, and nurses, ensuring confidentiality and interviewing participants at their preferred locations. Each questionnaire took approximately 15 minutes, while KIIs lasted 20–30 minutes. Data collection was conducted over a four-month period, from 10^th^ January 2023 to 19^th^ April 2023. This timeframe ensured adequate sampling to reflect diverse experiences across the selected counties. By integrating both quantitative and qualitative methodologies, this study provided a comprehensive understanding of the factors influencing neonatal weight gain in KMC settings.

### Data Management

Collected data underwent cleaning, coding, and entry to ensure completeness and consistency. Quantitative data was processed using SPSS version 26.0, with descriptive statistics (frequencies, means, and standard deviations) used for summarization. Bivariate analysis assessed differences in proportions, with p-values determining statistical significance. The Crude Odds Ratio (COR) and 95% Confidence Interval (CI) were used to measure the strength of associations between independent and dependent variables. Significant variables were further analyzed using binary logistic regression (enter method), where Adjusted Odds Ratios (AOR) and 95% CI determined final associations. Statistical significance was set at α = 0.05, with two-sided p-values reported.

Qualitative data from KIIs was manually cleaned, coded, and analyzed thematically based on emerging patterns. The findings were reported in narrative form and integrated with quantitative results to provide deeper insights.

### Ethical Considerations

The study adhered to ethical standards, ensuring informed consent was obtained from all participants, with an emphasis on voluntary participation and confidentiality. Ethical approval was granted by Masinde Muliro University of Science and Technology (MMUST/IERC/105/2022) and the National Commission for Science, Technology, and Innovation (NACOSTI/P/22/21399). Permission to conduct the study was obtained from the county governments of Busia, Kisii, and Migori, as well as the participating health facilities. The study aimed to provide valuable insights into KMC practices and their effectiveness in resource-limited settings, contributing to improved neonatal care.

## Results

### Participants Characteristics

A majority of participants were over 30 years old, with 160 mothers (58.18%) falling into this age group, while 115 mothers (41.82%) were 30 years old or younger as shown in Table 2. In terms of education, the distribution was diverse, with 24 mothers (8.73%) having primary education, 76 (27.64%) having secondary education, 99 (36.00%) having college education, and the remaining 76 (27.64%) having attained a university degree. Marital status among the participants varied, with 31.27% being married, 34.91% cohabiting 26.91% single, and 6.91% widowed. The average number of other children among the mothers was 2.95 ±1.54, with 185 (67.27%) having three or fewer children and 90 (32.73%) having more than three children. Occupationally, the participants were engaged in various roles, including employment (25.45%), business (30.91%), farming (14.55%), and homemaking (29.09%). In terms of newborn characteristics, there were slightly more female infants (58.91%) than males (41.09%). The average birth weight of the newborns was 1578.85 ±232.72 grams, with 113 (41.09%) weighing equal to or below 1500 grams and 162 (58.91%) weighing above 1500 grams. Additionally, the average gestational age at birth was 35.59 ±2.30 weeks, with 146 infants (53.09%) born at or before 35 weeks’ gestation and 129 (46.91%) born after 35 weeks.

**Table 1:** Proportionate Sample Allocation for Quantitative Sampling

| Health Facility | Study Population | Sample Size |
| --- | --- | --- |
| Migori County Referral Hospital | 150 | 58 |
| Busia County Referral Hospital | 123 | 48 |
| Kisii County Referral Hospital | 70 | 27 |
| Nyamache Subcounty Hospital (Kisii) | 45 | 18 |
| Rongo Subcounty Hospital (Migori) | 96 | 37 |
| Awendo Subcounty Hospital (Migori) | 74 | 29 |
| Nambale Subcounty Hospital (Busia) | 72 | 28 |
| Khunyangu Subcounty Hospital (Busia) | 77 | 30 |
| **Total** | **707** | **275** |

**Table 2:** Summary of maternal and newborn sociodemographic aspects.

| Characteristics | Grouping | Frequency | Percent |
| --- | --- | --- | --- |
| Maternal Age | ≤30 Years | 115 | 41.82 |
|  | > 30 Years | 160 | 58.18 |
| Maternal Level of education | Primary | 24 | 8.73 |
|  | Secondary | 76 | 27.64 |
|  | College | 99 | 36.00 |
|  | University | 76 | 27.64 |
| Marital status | Single | 74 | 26.91 |
|  | Married | 86 | 31.27 |
|  | Widowed | 19 | 6.91 |
|  | Cohabiting | 96 | 34.91 |
| Number of other children | ≤3 | 185 | 67.27 |
|  | >3 | 90 | 32.73 |
| Maternal occupation | Employed | 70 | 25.45 |
|  | Business | 85 | 30.91 |
|  | Farmer | 40 | 14.55 |
|  | Housewife | 80 | 29.09 |
| Information source on KMC | Radio/TV | 68 | 24.73 |
|  | Phone Text/ Internet | 41 | 14.91 |
|  | Healthcare worker | 114 | 41.45 |
|  | Family and friends | 52 | 18.91 |
| Number people in the household | ≤5 | 197 | 71.64 |
|  | >5 | 78 | 28.36 |
| Monthly income | Below KES 5000 | 51 | 18.55 |
|  | KES 5000 - 10000 | 72 | 26.18 |
|  | KES 10000 - 15000 | 51 | 18.55 |
|  | KES 15000 - 20000 | 51 | 18.55 |
|  | Above KES 20000 | 50 | 18.18 |
| Newborn's sex | Male | 113 | 41.09 |
|  | Female | 162 | 58.91 |
| Newborn's Birth Weight | ≤1500 grams | 113 | 41.09 |
|  | > 1500 grams | 162 | 58.91 |
| Gestation at time of birth | ≤35 weeks | 146 | 53.09 |
|  | > 35 weeks | 129 | 46.91 |

*Descriptive analysis of maternal and newborn sociodemographic characteristics. Maternal age mean =32.28 ±6.20 years, Average number of other children =2.95 ±1.54, average number of people in the household =4.59 ±1.64, average weight of the newborn =1578.85 ±232.72 grams, and average gestational age of the newborn =35.59 ±2.30 weeks.*

### Health worker inputs to KMC implementation

Table 3 provides insights into health worker inputs to KMC implementation, as perceived by mothers participating in the study. The perception of health worker support in the practice of KMC varied among respondents. Approximately 23.64% rated the support as excellent, while 19.64% found it somewhat excellent. Conversely, 17.82% rated the support as not excellent, with 19.64% considering it somewhat not excellent. Regarding the timing of KMC initiation after delivery, responses were diverse. While 31.27% reported immediate initiation, significant proportions initiated KMC at 1 hour (20.73%), 2 hours (18.18%), and 3 hours (16.73%) post-delivery. Approximately 13.09% initiated KMC more than 3 hours after delivery. The satisfaction with the quality of KMC support received varied among respondents, with 25.82% being very satisfied and 22.91% satisfied. Conversely, 19.64% were very dissatisfied, and 15.64% were dissatisfied with the quality of care received. A majority (64.00%) reported receiving support from nurse, indicating a significant level of involvement from healthcare professionals in the KMC process. Participants’ satisfaction with the level of knowledge of healthcare workers (HCWs) on KMC showed that 33.09% being very satisfied and 17.45% being very dissatisfied. More than half of the respondents (59.27%) felt that HCWs addressed their concerns on KMC adequately, 40.73% felt otherwise. Similarly, 52.36% reported that HCWs provided feedback on the progress of the baby, indicating active involvement and communication from healthcare professionals. Lastly, respondents rated the readiness of HCWs to help with their KMC roles, with 54.18% considering HCWs more than somewhat ready and 14.91% and 14.18% considering them somewhat not ready and not ready, respectively.

**Table 3:** Rating of health workers’ inputs to KMC services by the caretakers.

| Health worker KMC support aspect | Grouping | Frequency | Percent |
| --- | --- | --- | --- |
| Rate health worker support in your practice of KMC | Excellent | 65 | 23.64 |
|  | Somewhat excellent | 54 | 19.64 |
|  | Neutral | 53 | 19.27 |
|  | Somewhat not excellent | 54 | 19.64 |
|  | Not excellent | 49 | 17.82 |
| How soon KMC initiated after delivery | Immediately | 86 | 31.27 |
|  | 1 Hour | 57 | 20.73 |
|  | 2 hours | 50 | 18.18 |
|  | 3 hours | 46 | 16.73 |
|  | > 3 hours | 36 | 13.09 |
| Satisfaction with quality of KMC | Very satisfied | 71 | 25.82 |
|  | Satisfied | 63 | 22.91 |
|  | Neutral | 44 | 16.00 |
|  | Dissatisfied | 43 | 15.64 |
|  | Very dissatisfied | 54 | 19.64 |
| Received any support from nurses | Yes | 176 | 64.00 |
|  | No | 99 | 36.00 |
| Satisfaction with level of knowledge of HCW on KMC | Very satisfied | 91 | 33.09 |
|  | Satisfied | 51 | 18.55 |
|  | Neutral | 42 | 15.27 |
|  | Dissatisfied | 43 | 15.64 |
|  | Very dissatisfied | 48 | 17.45 |
| HCWs address my concerns on KMC adequately | Yes | 163 | 59.27 |
|  | No | 112 | 40.73 |
| Rating readiness of HCW to help with my KMC roles | Ready | 91 | 33.09 |
|  | Somewhat ready | 58 | 21.09 |
|  | Neutral | 46 | 16.73 |
|  | Somewhat not ready | 41 | 14.91 |
|  | Not ready | 39 | 14.18 |
| HCW provides feedback on progress of the baby | Yes | 144 | 52.36 |
|  | No | 131 | 47.64 |

### Health worker roles in KMC

Nurses roles were identified from the key informant interviews. The roles range from clinical, educational, research, advocacy, and community engagement as summarized under each of the themes derived from the KII.

### Clinical implementation roles

Nurses described roles involving direct patient care, support, and coordination within the KMC unit. They play crucial roles in providing direct care to mothers and babies during KMC sessions, ensuring their comfort and well-being throughout the process as expressed by one of the nurses.

*"As a bedside nurse in the KMC unit, my role is to provide direct care to mothers and babies during KMC sessions."*.

Nurses also take up the role of a KMC coordinator thus overseeing all aspects of KMC care, from scheduling sessions to guaranteeing the availability of essential resources, thereby facilitating the smooth operation of KMC units as echoed by one nurse.

*“…As a KMC coordinator, my role is to coordinate all aspects of KMC care, from scheduling sessions to ensuring that all necessary resources are available."*

### Educational roles

Nurses emphasized roles related to educating and training both staff and parents about KMC. These roles are also paramount, as highlighted by nurses such as two nurses.

*“ In my role as a KMC educator, I conduct training sessions for both staff and parents on the principles and practices of KMC.” “…I serve as a resource nurse for KMC, providing support and guidance to both staff and families on KMC best practices."*

In their capacities as KMC educators and resource nurses, respectively, they conduct training sessions, equipping both staff and parents with the knowledge and skills necessary for effective KMC implementation and adherence.

### Research and quality improvement roles

Research and quality improvement initiatives are spearheaded by nurses. Through their roles, they engage in research endeavors, data collection, and clinical trials, all aimed at evaluating and enhancing the effectiveness of KMC practices as demonstrated by two nurses

*"…My role involves conducting research and quality improvement activities related to KMC…”* and "*…, I participate in clinical trials and studies to evaluate the effectiveness of KMC."*

### Advocacy and policy development roles

Advocacy and policy development are integral components of the KMC landscape, championed by nurses such as one who says that

*…"In my role as a KMC advocate, I work to raise awareness of KMC among healthcare professionals, policymakers, and the general public."*

Equally, as KMC advocates and policy developers, they endeavor to raise awareness of KMC’s significance among healthcare professionals and policymakers while shaping and implementing policies supportive of KMC integration within healthcare systems as reiterated by one interviewee

*"I’m responsible for developing and implementing policies and procedures related to KMC within our hospital."*

### Community engagement roles

Nurses mentioned roles involving communication with families, discharge planning, and community engagement. Acting as family liaison nurses and coordinators, they facilitate communication between families and healthcare teams, coordinate discharge planning, and organize community-based KMC training programs, thus fostering a supportive environment conducive to KMC’s success beyond hospital settings as demonstrated by two interviewees. *“..as a family liaison nurse in the KMC unit, my role is to facilitate communication between families and the healthcare team…"* and *"My role involves coordinating*

*KMC training programs for healthcare professionals both within our hospital and in the community."*

### Staff capacity on KMC

Thematic analysis of key informant interviews identified staff capacity in terms of formal training, practical experience, role of training in eliciting passion and improving advocacy skills. Those who did not have formal training on KMC thought it is important in improving their KMC support capacity.

### Formal training impact

Nurses recognized the value of formal training sessions in enhancing their understanding and practice of KMC. One nurse reflected on how training equipped her with up-to-date techniques and insights, enabling more informed and compassionate care provision.

*"The training equipped me with the latest techniques and insights into the benefits of KMC… Since then, I’ve been able to provide more informed and compassionate care."*

Similarly, others noted increased advocacy efforts post-seminar attendance, emphasizing the positive influence of such educational opportunities.

*"I attended a seminar on KMC last year… Since then, I’ve been more proactive in advocating for KMC among our patients and colleagues…" and "It was a great refresher on the latest guidelines and best practices… I’ve been more confident in my ability to provide support to mothers practicing KMC."*

### Practical experience

Many nurses emphasized the importance of practical experience alongside formal training with one interviewee highlighting the transformative effect of learning by doing, leading to improvements in KMC practice

*"Learning by doing made a huge difference… Since then, I’ve been practicing KMC regularly, and I’ve noticed significant improvements."*.

Others, found practical experience in the hospital setting to be invaluable, complementing the foundational knowledge gained from formal training

*"….while it laid a good foundation, I’ve found that practical experience in the hospital setting has been invaluable…. since then, I’ve been actively involved in KMC sessions, guiding mothers through the process and addressing any concerns they may have."*

### Lack of training

Some nurses expressed a lack of formal training opportunities and highlighted the need for continuous education. Three nurses voiced their eagerness to attend workshops or seminars in the future, recognizing the importance of ongoing learning

*"Unfortunately, I haven’t had the opportunity to receive any formal training on KMC… I’m eager to attend a workshop or seminar in the future…", "No, I haven’t received any formal training on KMC since I started*

Despite this, nurses remained committed to providing support to KMC mothers through self-initiated learning and on-the-job experiences.

*I strive to provide emotional support and encouragement to mothers practicing KMC…." and "Unfortunately, I haven’t had any formal training on KMC since I joined this unit… I’m committed to doing my best despite the lack of formal training."*

### Advocacy

Several nurses showed a passion for KMC and expressed a desire to advocate for its benefits. Some described increased involvement in KMC advocacy efforts, driven by the belief in its benefits. Their passion fueled advocacy efforts within their units and broader communities.

*"I’ve become more involved in advocating for KMC within our unit… I’m passionate about spreading awareness and knowledge about its benefits."*

*"It’s motivated me to become more actively involved in promoting KMC both within our hospital and in the wider community."*

*"While formal training is beneficial, I believe that practical experience is equally important in becoming proficient."*

### Continuous Learning

Nurses stressed the importance of continuous learning to stay updated with the latest practices in KMC. One nurse highlighted the refreshment of knowledge through periodic updates,

*"It was nice to refresh my knowledge and learn about updates or advancements… I make sure to provide them with all the support and information they require…"*

Another reiterated the impact of having a grounding in the fundamental KMC principles. *"While it was a while ago, the fundamentals of KMC have stayed with me… I’m constantly striving to improve my skills."*

### Health Facility KMC support

The findings from Table 4 provide insights into the infrastructure and support available for KMC within healthcare facilities. A majority of facilities reported having breast pumps (61.09%), milk banks (59.27%), weighing scales (56.36%), and radiant heaters (62.55%). However, fewer facilities had weight charts (52.73%), warm water facilities (58.91%), and dedicated spaces for KMC (58.55%). Regarding maternal support, just under half of the facilities reported providing adequate food for mothers (48.73%) and having seats available for mothers (61.82%). Similarly, approximately half of the facilities had rooms designated for breast milk expression (48.73%), while slightly more than half provided milk preparation equipment (57.09%). Hygiene and safety considerations were also assessed, with 60.00% of facilities promoting hygiene and 56.36% guaranteeing safety. Additionally, a majority of facilities displayed adequate information on KMC (60.73%) and ensured that the information displayed was easy to read and understand (51.27%). Hospital management involvement comprised hospital manager checking on the KMC mothers and outreach programs. Approximately half of the facilities reported hospital managers checking on KMC mothers (53.82%) and having outreach programs for KMC (50.18%). Moreover, a similar proportion of hospitals supported family-centered KMC (57.45%) and had adequate staff to support KMC (61.82%). The average facility implementation score was 10.78 ±2.59 out of a possible score of 19, with a percent facility implementation of 56.71 ±13.65%.

**Table 4:** Facility level availability of resources for implementation of KMC.

| Facility implementation characteristics | Grouping | Frequency | Percent |
| --- | --- | --- | --- |
| Breast pump | Yes | 168 | 61.09 |
|  | No | 107 | 38.91 |
| Milk bank | Yes | 163 | 59.27 |
|  | No | 112 | 40.73 |
| Weighing scale | Yes | 155 | 56.36 |
|  | No | 120 | 43.64 |
| Weight chart | Yes | 145 | 52.73 |
|  | No | 130 | 47.27 |
| Warm water | Yes | 162 | 58.91 |
|  | No | 113 | 41.09 |
| Radiant heater | Yes | 172 | 62.55 |
|  | No | 103 | 37.45 |
| Space for KMC | Yes | 161 | 58.55 |
|  | No | 114 | 41.45 |
| Adequate food for the mother | Yes | 134 | 48.73 |
|  | No | 141 | 51.27 |
| Seats for the mother | Yes | 170 | 61.82 |
|  | No | 105 | 38.18 |
| Room for breast milk expression | Yes | 134 | 48.73 |
|  | No | 141 | 51.27 |
| Milk preparation equipment | Yes | 157 | 57.09 |
|  | No | 118 | 42.91 |
| Promotes hygiene | Yes | 165 | 60.00 |
|  | No | 110 | 40.00 |
| Guarantees safety | Yes | 155 | 56.36 |
|  | No | 120 | 43.64 |
| Adequate information on KMC displayed in the KMC spaces | Yes | 167 | 60.73 |
|  | No | 108 | 39.27 |
| Information displayed about KMC is easy to read and understand | Yes | 141 | 51.27 |
|  | No | 134 | 48.73 |
| Hospital managers check on KMC mothers | Yes | 148 | 53.82 |
|  | No | 127 | 46.18 |
| Hospital has outreach program for KMC | Yes | 138 | 50.18 |
|  | No | 137 | 49.82 |
| Hospital support family centered KMC | Yes | 158 | 57.45 |
|  | No | 117 | 42.55 |
| Hospital has adequate staff to support KMC | Yes | 170 | 61.82 |
|  | No | 105 | 38.18 |

*Descriptive summary of facility implementation of KMC. Overall implementation was $10.78 \pm 2.59$ out of a possible score of 19, Percent facility implementation $56.71 \pm 13.65$*

### Outcome of KMC for low birthweight babies in Hospitals, in Western Kenya

The frequency Table 5 provides comprehensive insights into various newborn outcome variables associated with KMC, offering a detailed understanding of the effectiveness of this care method in promoting the health and well-being of newborns. The birth weight distribution revealed that 113 newborns (41.09%) had a birth weight of less than or equal to 1500 grams, while 162 newborns (58.91%) had a birth weight exceeding 1500 grams. The average birth weight was 1578.85 ± 232.72 grams, and a range from 1202 to 2000 grams. At the time of data collection, 151 newborns (54.90%) exhibited a weight equal to or greater than 2000 grams, while 124 newborns (45.10%) had a weight less than 2000 grams. The average baby weight at data collection was 2010.12 ± 258.82 grams, and a range from 1299 to 2539 grams. In achieving the recommended daily weight gain, 105 newborns (38.18%) achieved the criterion of at least 15 grams per kilogram, while 170 newborns (61.82%) did not meet this target. The average daily weight gain at the time of data collection was 12.96 ±4.85 grams, and a range from 5 to 21 grams. Regarding temperature, 129 newborns (46.91%) had temperatures within the normal range at the time of data collection, while 146 newborns (53.09%) did not. The average temperature at data collection was 36.46 ± 0.83 degrees Celsius, and a range from 35.1 to 37.9 degrees Celsius. A majority of newborns, accounting for 198 (72.00%), exhibited a respiratory rate within the normal range, while 77 newborns (28.00%) did not. The average respiratory rate at data collection was 52.70 ± breaths per minute, and a range from 35 to 70 breaths per minute. Finally, the number of days spent on KMC compared to the recommended length of stay to achieve appropriate weight gain based on birth weight, 151 newborns (54.91%) were within the recommended length of stay, while 124 newborns (45.09%) did not. The average number of days on KMC was 36.25 days ± 10.09 days, and a range from 20 to 61 days. The recommended length of stay based on birth weight to attain appropriate weight gain had an average of 41.21 ± 16.02 days, and a range from 17 to 72 days.

**Table 5:**
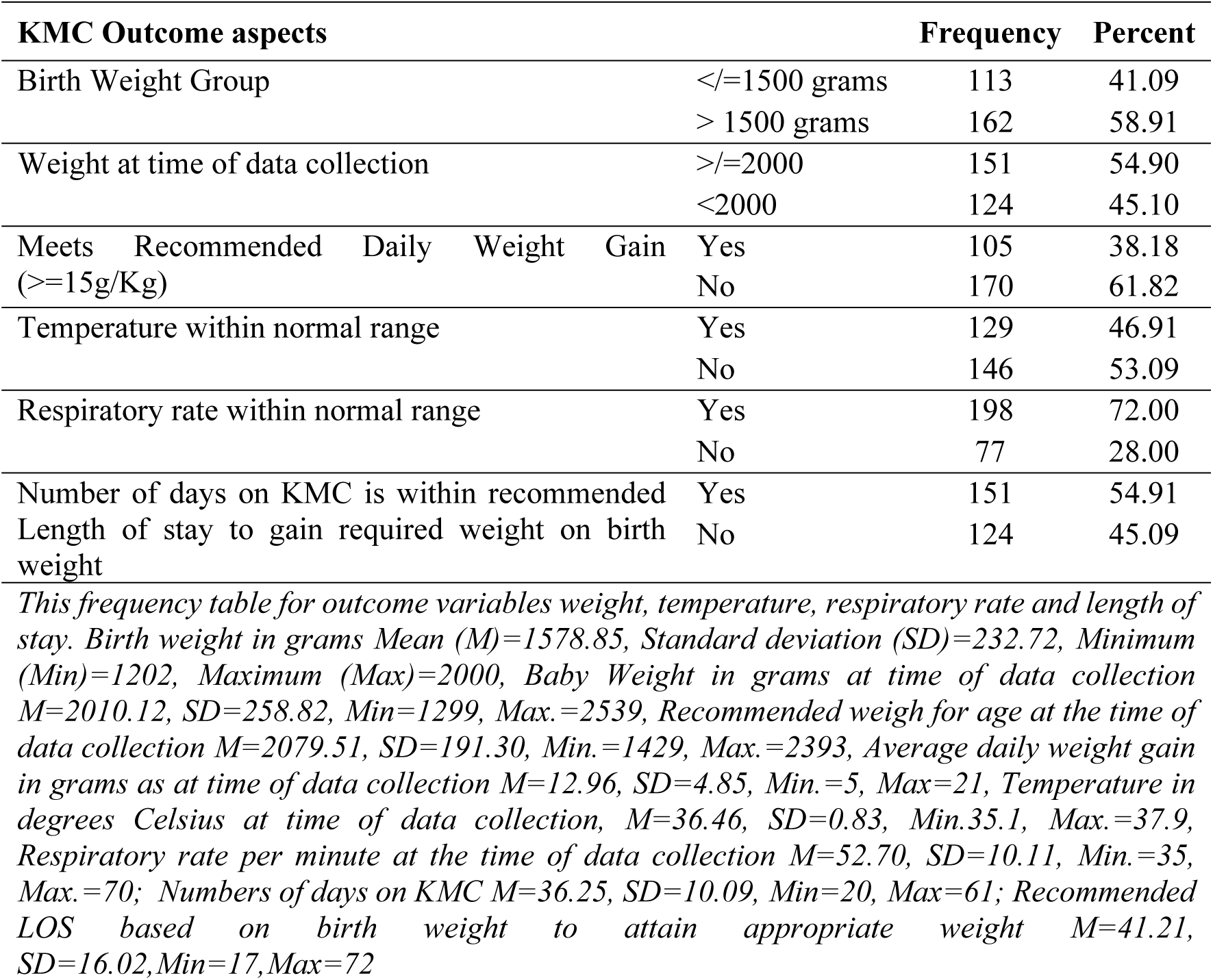
Outcome of KMC for low birthweight babies in Hospitals, in Western Kenya.

| KMC Outcome aspects |  |  |  |  | Frequency | Percent |
| --- | --- | --- | --- | --- | --- | --- |
| Birth Weight Group | </=1500 grams |  |  |  | 113 | 41.09 |
|  | > 1500 grams |  |  |  | 162 | 58.91 |
| Weight at time of data collection | >/=2000 |  |  |  | 151 | 54.90 |
|  | <2000 |  |  |  | 124 | 45.10 |
| Meets Recommended Daily Weight Gain (≥15g/Kg) | Yes | | | | 105 | 38.18 |
|  | No |  |  |  | 170 | 61.82 |
| Temperature within normal range | Yes |  |  |  | 129 | 46.91 |
|  | No |  |  |  | 146 | 53.09 |
| Respiratory rate within normal range | Yes |  |  |  | 198 | 72.00 |
|  | No |  |  |  | 77 | 28.00 |
| Number of days on KMC is within recommended Length of stay to gain required weight on birth weight | Yes |  |  |  | 151 | 54.91 |
|  | No |  |  |  | 124 | 45.09 |

*This frequency table for outcome variables weight, temperature, respiratory rate and length of stay. Birth weight in grams Mean (M)=1578.85, Standard deviation (SD)=232.72, Minimum (Min)=1202, Maximum (Max)=2000, Baby Weight in grams at time of data collection M=2010.12, SD=258.82, Min=1299, Max.=2539, Recommended weigh for age at the time of data collection M=2079.51, SD=191.30, Min.=1429, Max.=2393, Average daily weight gain in grams as at time of data collection M=12.96, SD=4.85, Min.=5, Max=21, Temperature in degrees Celsius at time of data collection, M=36.46, SD=0.83, Min.35.1, Max.=37.9, Respiratory rate per minute at the time of data collection M=52.70, SD=10.11, Min.=35, Max.=70; Numbers of days on KMC M=36.25, SD=10.09, Min=20, Max=61; Recommended LOS based on birth weight to attain appropriate weight M=41.21, SD=16.02,Min=17,Max=72*

### Health facility factors influencing neonatal weight gain under KMC in Western Kenya

Binary regression analysis was used to examine the association between health facility-related factors and the likelihood of neonates achieving the recommended daily weight gain of at least 15g/kg in Kangaroo Mother Care (KMC) settings. The findings presented in Table 6 indicate that several facility-level interventions significantly influence neonatal growth outcomes. The logistic regression model demonstrated good explanatory power, with a Nagelkerke R² of 0.605, suggesting that the model accounted for approximately 60.5% of the variance in weight gain outcomes. The Hosmer and Lemeshow goodness-of-fit test yielded a χ² value of 6.24 (df = 8, P = 0.620), indicating that the model fit the data well. After adjusting for potential confounders, eight facility-related factors were significantly associated with achieving the recommended weight gain. The presence of a breast pump (AOR: 2.81, 95% CI: 1.30–6.08, P = 0.009) and a milk bank (AOR: 3.22, 95% CI: 1.47–7.06, P = 0.004) significantly increased the likelihood of infants meeting the daily weight gain threshold. Similarly, provision of adequate food for mothers was one of the strongest predictors, with infants whose mothers received sufficient nutrition being 5.32 times more likely to gain weight adequately (AOR: 5.32, 95% CI: 2.45–11.55, P < 0.001). The availability of seating for mothers (AOR: 3.72, 95% CI: 1.68–8.28, P = 0.001) and a dedicated room for breast milk expression (AOR: 4.77, 95% CI: 2.22–10.23, P < 0.001) were also strong predictors of weight gain, highlighting the importance of providing comfortable and private spaces for maternal care in KMC facilities. Additionally, facilities that displayed easily understandable information about KMC (AOR: 2.86, 95% CI: 1.33–6.17, P = 0.007) significantly improved neonatal weight gain outcomes. Moreover, family-centered KMC support was an important factor, with hospitals that encouraged family involvement increasing the likelihood of adequate weight gain (AOR: 2.41, 95% CI: 1.08–5.40, P = 0.032). Finally, adequate staffing to support KMC emerged as a critical determinant, with infants in well-staffed facilities being 3.44 times more likely to meet the weight gain target (AOR: 3.44, 95% CI: 1.59–7.45, P = 0.002).

**Table 6:** Binary Regression Analysis of Health Facility Factors Associated with Neonatal Weight Gain in KMC.

| Health Facility related factors | | Daily Weight Gain $\geq 15\text{g/Kg}$ | | Bivariate Analysis | Binary Regression Analysis |
| --- | --- | --- | --- | --- | --- |
|  |  | Yes N(%) | No N(%) | COR [95% CI] P | B {COR [95% CI] P} |
| Breast pump | Yes | 80 (47.62) | 88 (52.38) | 2.98 [1.74-5.12]<br>**<0.001** | 1.03 {2.81 [1.30-6.08]<br>**0.009**} |
|  | No | 25 (23.36) | 82 (76.64) |  |  |
| Milk bank | Yes | 73 (44.79) | 90 (55.21) | 2.03 [1.21-3.39] | 1.17 {3.22 [1.47-7.06]} |
|  | No | 32 (28.57) | 80 (71.43) | **0.008** | **0.004** |
| Weighing scale | Yes | 62 (40.00) | 93 (60.00) | 1.19 [0.73-1.95] | 0.60{1.82 [0.84-3.93] |
|  | No | 43 (35.83) | 77 (64.17) | 0.532 | 0.130} |
| Weight chart | Yes | 59 (40.69) | 86 (59.31) | 1.25 [0.77-2.04] | -0.12{0.89 [0.43-1.85] |
|  | No | 46 (35.38) | 84 (64.62) | 0.386 | 0.757} |
| Warm water | Yes | 69 (42.59) | 93 (57.41) | 1.59 [0.96-2.63] | 0.70{2.02 [0.92-4.42] |
|  | No | 36 (31.86) | 77 (68.14) | 0.078 | 0.080} |
| Radiant heater | Yes | 75 (43.60) | 97 (56.40) | 1.88 [1.12-3.17] | 0.77{2.15 [1.00-4.63] |
|  | No | 30 (29.13) | 73 (70.87) | **0.021** | 0.051} |
| Space for KMC | Yes | 70 (43.48) | 91 (56.52) | 1.74 [1.05-2.88] | 0.76{2.13 [0.96-4.71] |
|  | No | 35 (30.70) | 79 (69.30) | **0.033** | 0.062} |
| Adequate food for the mother | Yes | 75 (55.97) | 59 (44.03) | 4.7 [2.77-7.98] | 1.67{5.32 [2.45-11.55] |
|  | No | 30 (21.28) | 111 (78.72) | **<0.001** | **<0.001** |
| Seats for the mother | Yes | 79 (46.47) | 91 (53.53) | 2.64 [1.54-4.51] | 1.32{3.72 [1.68-8.28] |
|  | No | 26 (24.76) | 79 (75.24) | **<0.001** | **0.001** |
| Room for breast milk expression | Yes | 70 (52.24) | 64 (47.76) | 3.31 [1.99-5.52] | 1.56{4.77 [2.22-10.23] |
|  | No | 35 (24.82) | 106 (75.18) | **<0.001** | **<0.001** |
| Milk preparation equipment | Yes | 70 (44.59) | 87 (55.41) | 1.91 [1.15-3.16] | -0.08{0.92 [0.42-2.00] |
|  | No | 35 (29.66) | 83 (70.34) | **0.012** | 0.834} |
| Promotes hygiene | Yes | 77 (46.67) | 88 (53.33) | 2.56 [1.51-4.34] | 0.69{2 [0.94-4.24] |
|  | No | 28 (25.45) | 82 (74.55) | **<0.001** | 0.0720} |
| Guarantees safety | Yes | 64 (41.29) | 91 (58.71) | 1.36 [0.83-2.22] | 0.17{1.18 [0.56-2.49] |
|  | No | 41 (34.17) | 79 (65.83) | 0.261 | 0.656} |
| Adequate information on KMC is displayed | Yes | 75 (44.91) | 92 (55.09) | 2.12 [1.26-3.56] | 0.03{1.03 [0.48-2.20] |
|  | No | 30 (27.78) | 78 (72.22) | **0.005** | 0.944} |
| KMC Information displayed is Simple | Yes | 66 (46.81) | 75 (53.19) | 2.14 [1.30-3.53] | 1.05{2.86 [1.33-6.17] |
|  | No | 39 (29.10) | 95 (70.90) | **0.003** | **0.007** |
| Hospital managers check on KMC mothers | Yes | 65 (43.92) | 83 (56.08) | 1.7 [1.04-2.80] | 0.50{1.65 [0.76-3.60] |
|  | No | 40 (31.50) | 87 (68.50) | **0.046** | 0.206} |
| Hospital has outreach program for KMC | Yes | 65 (47.10) | 73 (52.90) | 2.16 [1.31-3.55] | 0.47{1.6 [0.73-3.48] |
|  | No | 40 (29.20) | 97 (70.80) | **0.003** | 0.237} |
| Hospital support family cantered KMC | Yes | 70 (44.30) | 88 (55.70) | 1.86 [1.12-3.09] | 0.88{2.41 [1.08-5.40] |
|  | No | 35 (29.91) | 82 (70.09) | **0.017** | **0.032** |
| Hospital has adequate staff to support KMC | Yes | 79 (46.47) | 91 (53.53) | 2.64 [1.54-4.51] | 1.24{3.44 [1.59-7.45] |
|  | No | 26 (24.76) | 79 (75.24) | **<0.001** | **0.002** |
| Level of facility implementation (%) | ≥50 | 102 (54.26) | 86 (45.74) | 33.2[10.1-108.8] | -1.23{0.29 [0.06-1.36] |
|  | <50 | 3 (3.45) | 84 (96.55) | **<0.001** | 0.117} |

*Bivariate analysis by cross-tabulation with Chi square statistic as the inferential statistics, N (%) where N = counts, and %= percentage, $\alpha \leq 0.05$ . Binary regression analysis, enter mode, Nagelkerke $R^2$ 0.605, Hosmer and Lemeshow Test (6.24,8)=0.620, COR-Crude Odds Ratio, AOR - Adjusted Odds Ratio, 95% CI -95% Confidence Interval.*

While several factors were significant in bivariate analysis, some did not retain their significance in the adjusted regression model. These included the availability of weighing scales, weight charts, warm water, radiant heaters, space for KMC, hygiene promotion, managerial oversight of KMC mothers, outreach programs, and overall facility implementation level. Despite showing initial associations, these factors were not independent predictors of neonatal weight gain after controlling for confounders.

### Nurses’ views on the role of the facility in the provision of KMC

Thematic analysis of the nurses’ responses regarding the role of the facility in the provision of Kangaroo Mother Care (KMC) revealed several key themes:

### Management Support and Leadership

Management Support and Leadership were emphasized as facilitators of effective implementation of KMC protocols. This support enables the allocation of resources, training of staff, and maintenance of necessary infrastructure for KMC units.

One respondent highlighted this, stating, *"The facility plays a crucial role in the provision of KMC. Firstly, the management support system sets the tone for implementing KMC protocols effectively*."

Strong leadership commitment at all levels of the facility was portrayed as essential for better implementation of KMC initiatives, leading to support and accountability, thus leading to sustainability.

This was highlighted by the sentiment that *“Leadership commitment is key. Strong leadership at all levels of the facility, from senior management to unit supervisors, is essential for driving the implementation of KMC initiatives, fostering a culture of support and accountability, and ensuring sustainability in the long run.”*

Equally the nurses emphasized the importance of;

*“regular supervision and feedback sessions to ensure adherence to KMC protocols and identify areas for improvement.”*

One respondent stated, *"Supervision is another critical factor."*

Provision of training opportunities was also highlighted as crucial for both new and existing staff to stay *“…updated with the latest techniques and guidelines related to KMC.”*

These two are management functions to enhance staff capacity and ensure continuous quality improvement. Equally, the nurses stressed the importance of financial support for implementing KMC protocols, including investments in equipment, training programs, and hiring additional staff. Clear guidelines and protocols were also deemed essential for ensuring “*consistency in care delivery and minimizing errors*.”

### Resource appropriation

Resource appropriation such as the availability of resources like kangaroo chairs, breastfeeding support equipment, and monitoring devices, emerged as a significant factor influencing KMC provision. One of the nurses mentioned that;

"*Resources are a significant aspect… Adequate provision of essential resources is essential for providing quality KMC*."

Additionally, nurses discussed how rational allocation of nurses directly impacts the provision of KMC, as a heavy workload or allocation of nurses who are not specialized in KMC can lead to;

*“…less time spent on each baby, affecting the quality of care provided.”*

### Community Engagement

Community Engagement was recognized as important external factors in supporting KMC provision through anecdotes like;

"… *external factors such as community engagement and support also play a role.".* Equally, it was stated that “…*understanding and respecting the cultural beliefs and practices of the mothers and families receiving KMC is crucial for building trust and rapport. It can influence their willingness to engage in KMC and follow through with the recommended practices.”*

### Infrastructure and Communication

Infrastructure, including the physical layout of the facility, was deemed crucial for promoting skin-to-skin contact between mother and baby. A nurse emphasized that,

*"Infrastructure is key. The physical layout of the facility should be designed in a way that promotes skin-to-skin contact between the mother and the baby. This includes comfortable spaces for mothers to practice KMC, privacy when needed, and easy accessibility to support services like lactation consultants. "*

Effective communication channels within the facility were also highlighted as essential for smooth coordination in providing KMC as it;

*“…facilitates the transfer of information about mothers and babies who need KMC, ensures timely interventions, and prevents any gaps in care.”*

### Maternal involvement in quality improvement

Maternal involvement in quality improvement was highlighted as key. The nurses discussed the importance of addressing barriers to maternal involvement in KMC and continuous quality improvement initiatives within the facility. One nurse highlighted the need to address barriers to maternal involvement, stating;

*"Addressing barriers to maternal involvement is crucial."* Equally, ***“r****esearch and data collection”* within the facility was recognized as beneficial for monitoring the effectiveness of the KMC program and identifying areas for continuous improvement.

## Discussion

This study demonstrates the critical role of facility support in KMC implementation and its impact on neonatal weight gain. While health care worker (HCW) inputs were generally positive, inconsistencies in timely KMC initiation and HCW readiness were noted. Nurses played diverse roles, including direct care, training, advocacy, research, and community engagement, yet staff capacity varied, with gaps in formal KMC training. Facility-level implementation revealed infrastructure inconsistencies, particularly in maternal support and dedicated KMC spaces, which may have influenced neonatal outcomes. The study found that only 38.18% of LBW infants achieved the recommended weight gain, with challenges such as temperature instability and respiratory complications persisting. Provision of breast pumps, milk banks, adequate maternal nutrition, adequate seating, dedicated lactation spaces, family-centered care, display of clear easy to understand health messages on KMC, and sufficient staffing were identified as key facility factors associated with improved weight gain. Nurses emphasized the facility’s role in leadership, supervision, training, and community engagement, highlighting the need for enhanced policies, improved infrastructure, and continuous capacity-building to optimize KMC effectiveness and improve neonatal survival outcomes.

Facility support is critical in KMC implementation and its impact on neonatal weight gain. Despite its benefits, only 38.18% of LBW infants achieved the recommended weight gain, with challenges such as temperature instability and respiratory complications persisting. These findings align with research showing that LBW infants are highly susceptible to thermoregulation issues, which can lead to hypothermia, apnea, and desaturation episodes [14, 15]. Growth faltering within the first six months remains a concern, particularly among preterm infants [16]. However, Suyami et al., 2023 demonstrated KMC effectiveness in stabilizing body temperature and improving growth, reinforcing the importance of maternal involvement in neonatal care [17].

The findings align with existing literature, demonstrating that while health worker contributions were generally positive, inconsistencies in timely KMC initiation and HCW readiness remain a concern. In Nyeri County, Kenya, 89% of HCWs held favorable views on KMC [18], though inconsistencies in KMC initiation and operational readiness remained. A Malawi survey found that while 79% of hospitals provided KMC services but only 43% met operational standards [19]. Similar gaps exist in Kenya, where only half of HCWs have received KMC training, with incubators still preferred in some facilities [18]. These findings emphasize the need for ongoing training and better resource allocation to ensure timely and effective KMC implementation and onboarding of staff to support the initiative [19, 20].

Nurses play a key role in direct patient care, training, advocacy, research, and community engagement in KMC, but gaps in formal KMC training persist. The On-Site Nurse Mentoring program in Bihar has shown that practical, hands-on training improves nursing competencies, particularly in neonatal care [21]. However, frequent staff transfers and lack of structured mentorship hinder continuity [22]. The Clinical Nurse Specialist role further expands educational and advocacy responsibilities, but engagement levels vary significantly [23]. The low-dose, high-frequency training model has been effective in building provider skills in low-resource settings, reinforcing the need for structured, continuous training in KMC [24].

At facility level, inconsistent infrastructure and maternal support significantly affect KMC outcomes. While KMC reduces neonatal mortality, its adoption varies across facilities, with gaps in adherence to national guidelines in Bangladesh and Uganda [25, 26]. In Malawi, improved neonatal ward infrastructure resulted in higher survival rates, emphasizing the role of adequate space and trained staff [27]. A study in India, found that only 61% of primary health centers met structural requirements for maternal and neonatal care, reflecting resource limitations that hinder quality of care for KMC [28]. The current study demonstrated that there is need for targeted investments in KMC facility infrastructure to improve neonatal outcomes.

This study identified several facility-level factors significantly contributing to neonatal weight gain in KMC settings, including breast pumps, milk banks, maternal nutrition, comfortable seating, dedicated lactation spaces, family-centered care, clear KMC information, and sufficient staffing. Access to breast pumps and milk banks is critical for neonatal weight gain. Research confirms that mother’s own milk is the optimal nutritional source for preterm and low birth weight (LBW) infants, providing essential immune protection, gut maturation support, and nutrient absorption [29]. Our findings align with these insights, showing that facilities with milk banks and breast pumps provided better nutritional support, enhancing neonatal weight gain outcomes.

Lactation support services, including dedicated breastfeeding spaces, lactation counseling, and readily accessible information, are crucial for improving breastfeeding initiation and continuity among mothers practicing KMC [29]. Proactive lactation support is linked to higher rates of exclusive breastfeeding, which directly correlates with improved neonatal weight gain and better long-term growth outcomes [30]. Creating a supportive environment extends to the physical comfort of mothers as well. Comfortable seating arrangements contribute to longer skin-to-skin contact durations, essential for infant thermoregulation, weight gain, and physiological stability. Research confirms that providing ergonomic chairs or resting areas enhances KMC effectiveness by allowing mothers to sustain prolonged skin-to-skin contact without physical strain [31]. Furthermore, access to clear, easy-to-understand KMC information positively influences maternal knowledge and practice. Facilities with structured, visually engaging, and culturally relevant educational materials on KMC demonstrate higher maternal engagement and better neonatal outcomes. This aligns with existing research showing that structured health education enhances KMC adherence and neonatal growth [30, 32]. Our findings demonstrate that facilities prioritizing key elements of a supportive KMC environment experience better neonatal weight gain. This shows the need for investment in well-equipped lactation spaces that are comfortable with access to KMC for better outcome.

Adequate maternal nutrition is another critical factor influencing neonatal weight gain. Malnourished mothers often experience low milk production, hindering optimal infant feeding. Providing nutritious meals for lactating mothers has been associated with improved milk supply, maternal well-being, and breastfeeding success [33, 34]. Our study supports this, showing that facilities offering adequate maternal nutrition had higher rates of neonatal weight gain compared to those with limited food support. It is, therefore, important to integrate maternal nutrition programs into neonatal and KMC services to improve neonatal growth outcomes.

Family-centered care significantly enhances maternal adherence to KMC. Facilities involving family members in KMC practices observed higher compliance rates, as mothers received emotional and physical support from partners and relatives. Studies indicate that family involvement reduces maternal stress and strengthens adherence, leading to better neonatal health outcomes [35–38].

Adequate staffing emerged as a key determinant of KMC success. Facilities with sufficiently trained healthcare workers were better equipped to support mothers, monitor infants, and ensure adherence to best KMC practices. In contrast, understaffed facilities struggled to provide consistent guidance, negatively affecting maternal engagement and neonatal outcomes. These findings are consistent with previous studies showing that staffing shortages in neonatal units limit healthcare workers’ ability to provide personalized support, ultimately impacting neonatal growth and survival [38–40]. Ensuring optimal nurse-to-patient ratios and continuous KMC training for staff is critical for sustaining high-quality neonatal care.

Strong leadership, policy integration, and structured supervision are critical for sustaining KMC implementation and optimizing its effectiveness. Our findings highlight that integrating KMC into neonatal care policies improves neonatal survival and weight gain outcomes [27, 41, 42]. However, challenges such as inconsistent leadership support, insufficient funding, and weak health system integration hinder progress, mirroring barriers reported in other studies [43]. A well-structured healthcare system with skilled staff and adequate resources is essential for improving neonatal care, particularly in developing countries where over 90% of neonatal deaths occur [27, 41]. Evidence from Uganda demonstrates that training and supervision can enhance health worker competency, leading to a 30% increase in facility deliveries and an 85% survival rate for preterm infants in KMC units [10]. Despite these successes, gaps in policy, coverage, and newborn health integration into national frameworks persist. Institutional commitment, stronger health system support, and continuous capacity-building are essential to address these challenges, scale up KMC implementation, and sustain its impact on neonatal outcomes [44].

This study demonstrates the importance of facility support in optimizing KMC effectiveness and neonatal health outcomes. Addressing the gaps is crucial in ensuring that KMC remains a sustainable and effective intervention for low birth weight and preterm infants. Our findings contribute to the growing evidence supporting facility-based interventions as a key strategy for improving neonatal survival and growth in resource-limited settings.

### Study limitations

The cross-sectional approach captured data at a single point in time, restricting the ability to establish causal relationships between facility factors and neonatal weight gain. Self-reported data from mothers and healthcare workers may have introduced recall and social desirability bias, affecting response accuracy. Equally, facility-based design limits the generalizability of findings to community or home-based KMC practices, further, potential selection bias may have overrepresented mothers highly engaged in KMC yet adherence and support structures in home settings may differ. Variability in infrastructure, staffing, and resources across study sites may have influenced findings, making comparisons challenging. Despite these limitations, the study provides valuable insights into facility-level factors influencing KMC outcomes, informing policy and resource allocation for improved neonatal care.

### Conclusion and recommendations

Facility support is fundamental to optimizing KMC and improving neonatal weight gain. By prioritizing KMC policies, staff training, and maternal education, healthcare facilities can significantly enhance neonatal outcomes. Strengthening infrastructure, supervision, and staff capacity will be key to ensuring widespread and effective KMC implementation.

To optimize KMC implementation and improve neonatal weight gain, healthcare facilities should enhance neonatal feeding support by investing in breast pumps, human milk banks, and lactation support services, including dedicated breastfeeding spaces. Maternal support and engagement should be strengthened through integrated maternal nutrition programs and comfortable seating arrangements to facilitate prolonged skin-to-skin contact and improve neonatal thermoregulation. Increasing trained staff in neonatal units, promoting family involvement, and ensuring the availability of clear, accessible KMC education materials will enhance facility staffing and family-centered care. Additionally, leadership and supervision should be reinforced to ensure effective policy implementation, resource allocation, and continuous quality improvement in KMC services.

## Data Availability

The data underlying the results presented in the study are available from: https://osf.io/bv9x4/?view_only=698e140567124ee68e8ded62b323f813

## Acknowledgements

Special gratitude goes to School of Nursing, Midwifery and Paramedical Science for guidance and support. Sincere gratitude to county department of education, County director of health, all administrators in the selected health facilities and respondents who took part to make this study successful.

## Abbreviations

KMC: Kangaroo Mother Care
HCW: Health Care Worker
KII: Key Informant interview
COR: Crude Odds Ratio
AOR: Adjusted Odds Ratio
CI: Confidence Interval
SPSS: Statistical Package for Social Sciences

## Author contribution

**SM:** conceptualization and design of the study, conducting the literature review, collecting and analysing data, interpreting the results, and writing the manuscript.

**ENM:** Review of the concept/study design, examination of data analysis and interpretation and writing of the manuscript.

**MK:** Review of the concept and study design, examination of data analysis and interpretation and review of manuscript writing.

**MSS:** Data analysis, manuscript drafting, and journal correspondence.

